# Evidence of unexplained discrepancies between planned and conducted statistical analyses: a review of randomized trials

**DOI:** 10.1101/2020.02.20.20025684

**Authors:** Suzie Cro, Gordon Forbes, Nicholas A Johnson, Brennan C Kahan

## Abstract

**Background:** Choosing or altering the planned statistical analysis approach after examination of trial data (often referred to as ‘p-hacking’) can bias results of randomized trials. However, the extent of this issue in practice is currently unclear. We conducted a review of published randomized trials to evaluate how often a pre-specified analysis approach is publicly available, and how often the planned analysis is changed.

**Methods:** A review of randomised trials published between January and April 2018 in six leading general medical journals. For each trial we established whether a pre-specified analysis approach was publicly available in a protocol or statistical analysis plan, and compared this to the trial publication.

**Results:** Overall, 89 of 101 eligible trials (88%) had a publicly available pre-specified analysis approach. Only 22/89 trials (25%) had no unexplained discrepancies between the pre-specified and conducted analysis. Fifty-four trials (61%) had one or more unexplained discrepancies, and in 13 trials (15%) it was impossible to ascertain whether any unexplained discrepancies occurred due to incomplete reporting of the statistical methods. Unexplained discrepancies were most common for the analysis model (n=31, 35%) and analysis population (n=28, 31%), followed by the use of covariates (n=23, 26%) and the approach for handling missing data (n=16, 18%). Many protocols or statistical analysis plans were dated after the trial had begun, so earlier discrepancies may have been missed.

**Conclusions:** Unexplained discrepancies in the statistical methods of randomized trials are common. Increased transparency is required for proper evaluation of results.

## Background

The results of a clinical trial depend upon the statistical methods used for analysis. For example, changing the analysis population or method of handling missing data can change the size of the estimated treatment effect or its standard error. In some instances these differences can be large and may affect interpretation of the trial (1–6). If investigators choose the method of analysis based on trial data in order to obtain more favourable results (often referred to as ‘p-hacking’), this can cause bias (7). Selective reporting has been identified previously, where outcomes with more favourable results are more likely to be reported than other outcomes (8–19). There is some evidence to suggest this may also be a concern for statistical analyses; pre-specification of the proposed methods is often poor, discrepancies between protocols and publications are common, and in some instances changes may have been made to obtain specific results (5, 8, 10, 13, 20–23).

Guidelines such as ICH-E9 (24) (International Conference for Harmonisation of Technical Requirements for Pharmaceuticals for Human Use), SPIRIT (25, 26) (Standard Protocol Items: Recommendations for Interventional Trials), and CONSORT(27) (Consolidated Standards of Reporting Trials) require investigators to pre-specify the principle features of their statistical analysis approach in the trial protocol, and report any changes in the trial report. This strategy can reduce bias from analysis being chosen based on trial data, and allows readers to assess whether inappropriate changes were made.

We conducted a review of trials published in general medical journals to evaluate how often a pre-specified analysis approach was publicly available, how often the planned analysis approach was changed, whether these changes were explained, and the reporting around the timing and blinding status of changes.

## Methods

### Search strategy

In this review, we examined randomized controlled trials published between January and April 2018 in six general high impact medical journals: Annals of Internal Medicine; The BMJ; Journal of the American Medical Association (JAMA); The Lancet; New England Journal of Medicine (NEJM); and PLOS Medicine. We searched for articles in PubMed with a publication type of “randomized controlled trial” or categorised with the MeSH term “random allocation,” or including the keyword “random*” in the title or abstract, restricted to the aforementioned included journals and publication period. The full search strategy is shown in Appendix 1 in Additional File 1 and was conducted July 2018.

### Eligibility

Articles were eligible for inclusion if they reported results from a phase 2-4 randomized trial in humans. Exclusion criteria were pilot or feasibility study, phase 1 trial, non-randomized study, secondary analysis of previously published trial, cost-effectiveness as the primary outcome, more than one trial reported in the article, results of an interim analysis, or if the protocol or SAP was not in English.

One author screened the title and abstract of each paper for eligibility. The full texts of these articles were then assessed independently by two reviewers to confirm eligibility. For all eligible studies, one author searched the main text, supplementary material, and references to identify whether a protocol and/or SAP was available.

### Data extraction

Data was extracted onto a pre-piloted standardised data extraction form by two reviewers independently. Disagreements were resolved by discussion, or by a third reviewer where disagreement could not be resolved. Where the trial publication referred to supplementary material, a SAP or protocol, the extractor referred to these documents.

We extracted data related to the primary analysis of the primary outcome from the trial publication. A single primary outcome was identified as follows; (a) if one outcome was listed as the primary we used this; (b) if no outcomes or multiple outcomes were listed as being primary we used the outcome that the sample size calculation was based on; and (c) if no sample size calculation was performed or sample size was calculated for multiple primary outcomes, we used the first clinical outcome listed in the objectives/outcomes section. We identified the primary analysis as follows; (a) if a single analysis strategy was used, or multiple strategies were used with one being identified as primary, we used this; (b) if multiple strategies were used without one being identified as primary, we used the first one presented in the results section.

For each article, we extracted general trial characteristics, whether protocols or SAPs were available, including the dates of these documents and, if available, the blinding status of trial statisticians. For articles with a protocol or SAP, we compared the method of analysis in the trial publication against the method specified in the earliest available protocol or SAP which included some information on the analysis of the primary outcome (referred to as the original analysis plan). We assessed the following four analysis elements: (i) analysis population (the set of participants included in the analysis, and which treatment group they were analysed in); (ii) the statistical analysis model; (iii) use of baseline covariates in the analysis; and (iv) the method for handling missing data. We chose these elements as they are specified in the SPIRIT guidelines, and have been used in previous reviews (5, 25).

We evaluated two types of discrepancies for each analysis element. The first, termed a ‘change’, occurred when the analysis element in the trial publication was different to that specified in the original analysis plan. The following examples would constitute changes: (a) if an intention-to-treat analysis population was originally specified, but a per-protocol analysis was used; (b) if the functional form of the statistical analysis model was changed, such as from a mixed-effects regression model to generalized estimating equations (GEE); (c) if the original analysis plan specified the analysis would not adjust for baseline covariates but the trial publication adjusted for one or more patient characteristic; or (d) if a complete case analysis was originally specified, but multiple imputation was used.

The second discrepancy, termed an ‘addition’, occurred when the original analysis plan gave the investigators flexibility to subjectively choose the final analysis method after seeing trial data. This could occur if the original analysis plan (i) contained insufficient information about the proposed analysis; or (ii) allowed the investigators to subjectively choose between multiple different potential analyses. The following examples would constitute additions: if the original analysis plan stated that (a) both a per-protocol and intention-to-treat analysis population would be used, without specifying which was the primary analysis (as investigators could then decide during final analysis which was the primary, based on which gave the most favourable result); (b) either parametric or non-parametric methods would be used depending on distributional assumptions, but did not define an objective criteria for assessing distributional assumptions (as the investigators could then present whichever method gave the most favourable result); (c) the analysis would adjust for important baseline covariates, but did not define how these covariates would be chosen (as investigators could choose during final analysis the set of covariates which gave the most favourable result); or (d) multiple imputation would be used, but did not define what the method of imputation would be, or what variables would be included in the imputation model (as this would allow the investigators to run several different imputation models during final analysis and present only the most favourable).

We classified each discrepancy as being ‘explained’ or ‘unexplained’. Discrepancies were classified as explained if they had been specified in a subsequent version of the protocol or SAP (with or without a justification or rationale for the discrepancy), or if the trial publication explained that an alteration to the pre-specified analysis approach had been made. Otherwise discrepancies were classified as unexplained.

### Outcomes

The main outcome measures were (i) the number of trials with a publicly available pre-specified analysis approach for the primary outcome (i.e. whether an original analysis plan was available in a protocol or a SAP); (ii) the number of trials with no unexplained discrepancies from the publicly available pre-specified analysis approach; and (iii) the total number of analysis elements for each trial with an unexplained discrepancy.

Secondary outcomes were, for each analysis element described earlier, (i) the number of trials with at least one unexplained discrepancy (either change or addition); (ii) the number of trials with at least one unexplained change; and (iii) the number of trials with at least one unexplained addition.

### Statistical methods

Outcomes were summarised descriptively using frequencies and percentages. We performed two pre-specified subgroup analyses, where we summarised outcomes separately according to trial funding status, and type of intervention. One post-hoc subgroup analysis was performed according to availability of a SAP.

All statistical analyses were performed using Stata version 15 (28).

## Results

### Search results and characteristics of included studies

Our search identified 197 articles, of which 101 were eligible (see Fig 1 and for a list of eligible trials Appendix 2 in Additional File 1). General trial characteristics are shown in Table 1.

**Table 1.**
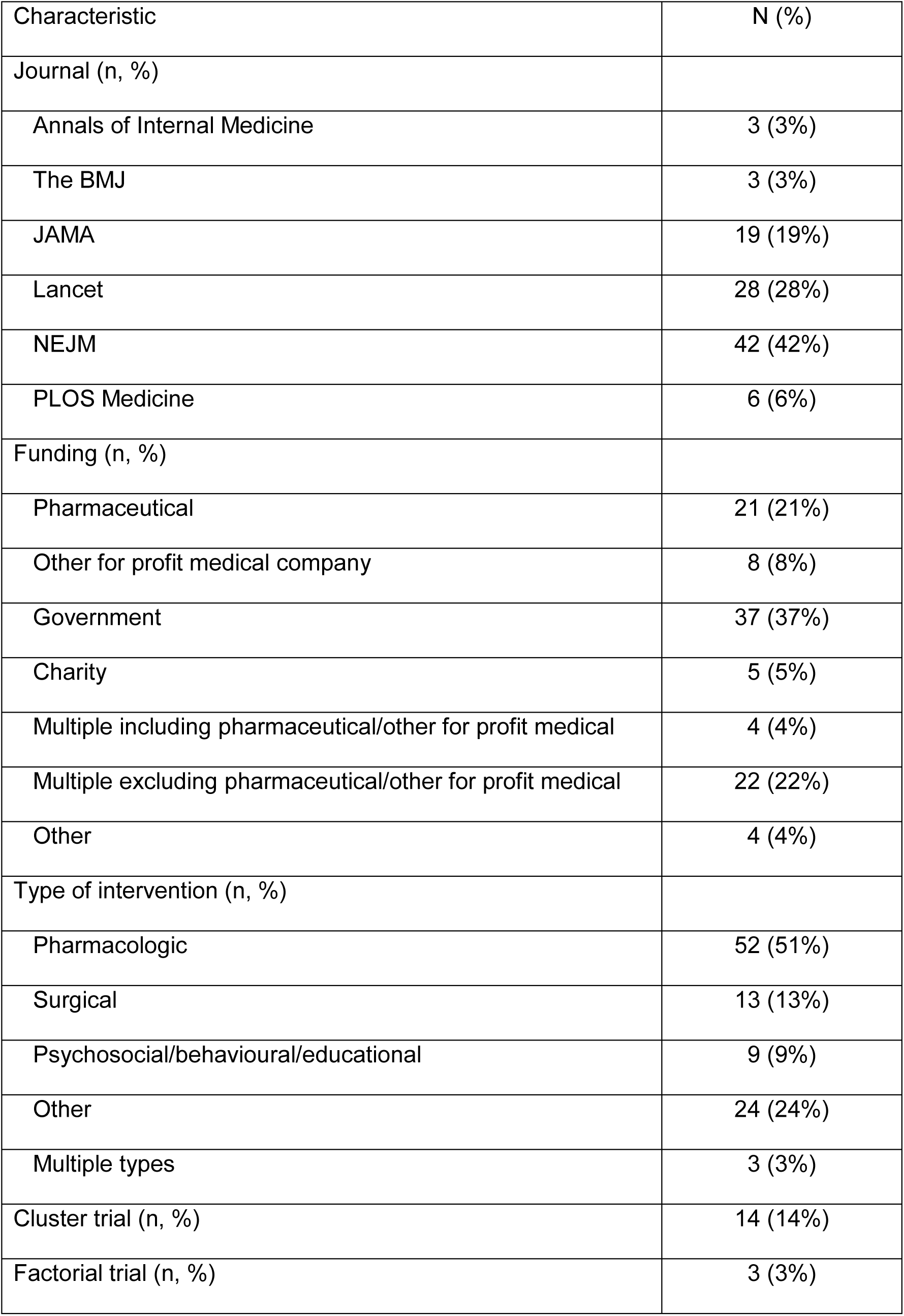
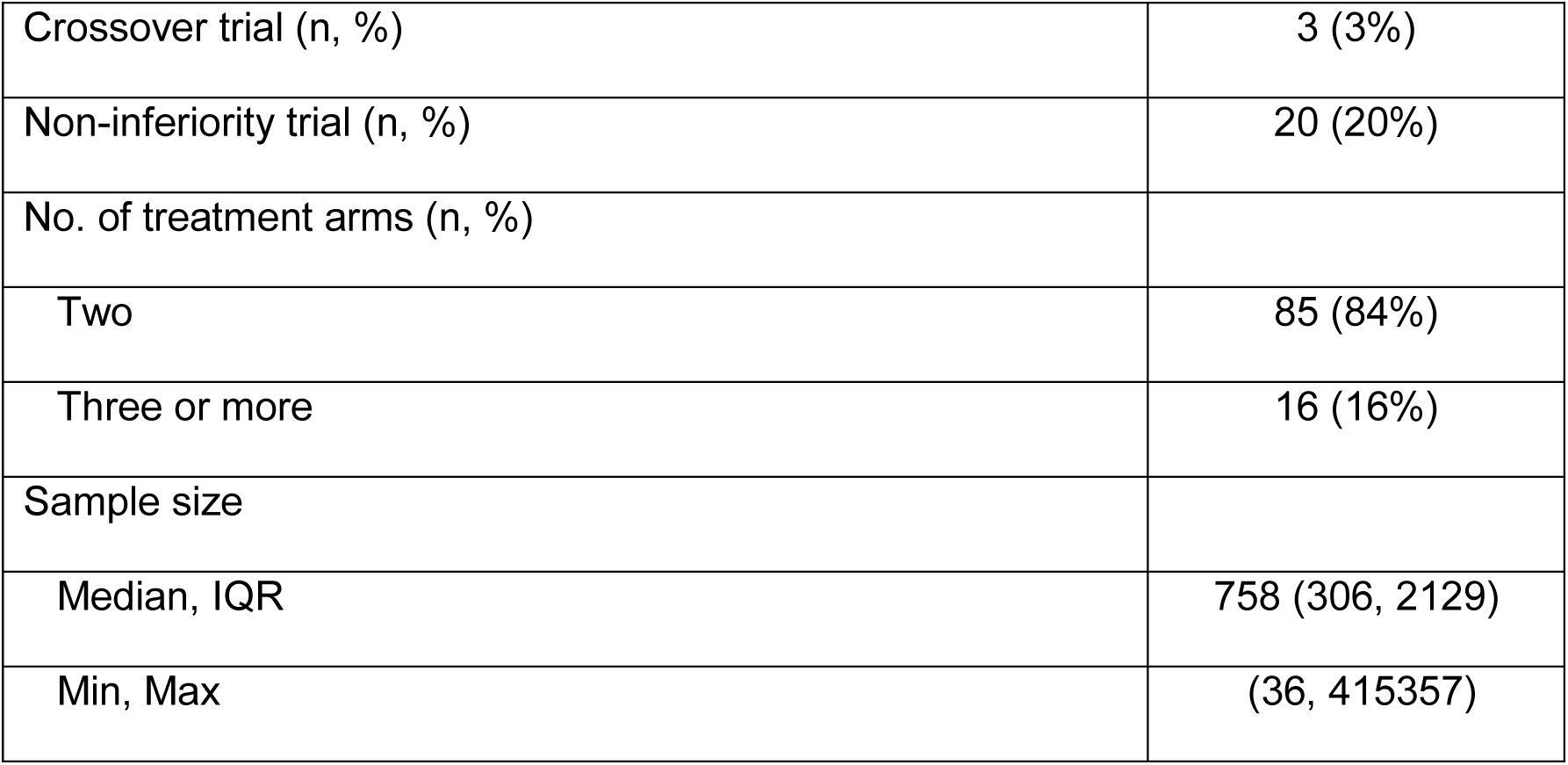
– Characteristics of eligible trials (N=101)

| Characteristic | N (%) |
| --- | --- |
| Journal (n, %) |  |
| Annals of Internal Medicine | 3 (3%) |
| The BMJ | 3 (3%) |
| JAMA | 19 (19%) |
| Lancet | 28 (28%) |
| NEJM | 42 (42%) |
| PLOS Medicine | 6 (6%) |
| Funding (n, %) |  |
| Pharmaceutical | 21 (21%) |
| Other for profit medical company | 8 (8%) |
| Government | 37 (37%) |
| Charity | 5 (5%) |
| Multiple including pharmaceutical/other for profit medical | 4 (4%) |
| Multiple excluding pharmaceutical/other for profit medical | 22 (22%) |
| Other | 4 (4%) |
| Type of intervention (n, %) |  |
| Pharmacologic | 52 (51%) |
| Surgical | 13 (13%) |
| Psychosocial/behavioural/educational | 9 (9%) |
| Other | 24 (24%) |
| Multiple types | 3 (3%) |
| Cluster trial (n, %) | 14 (14%) |
| Factorial trial (n, %) | 3 (3%) |
| Crossover trial (n, %) | 3 (3%) |
| Non-inferiority trial (n, %) | 20 (20%) |
| No. of treatment arms (n, %) |  |
| Two | 85 (84%) |
| Three or more | 16 (16%) |
| Sample size |  |
| Median, IQR | 758 (306, 2129) |
| Min, Max | (36, 415357) |

**Fig 1:**
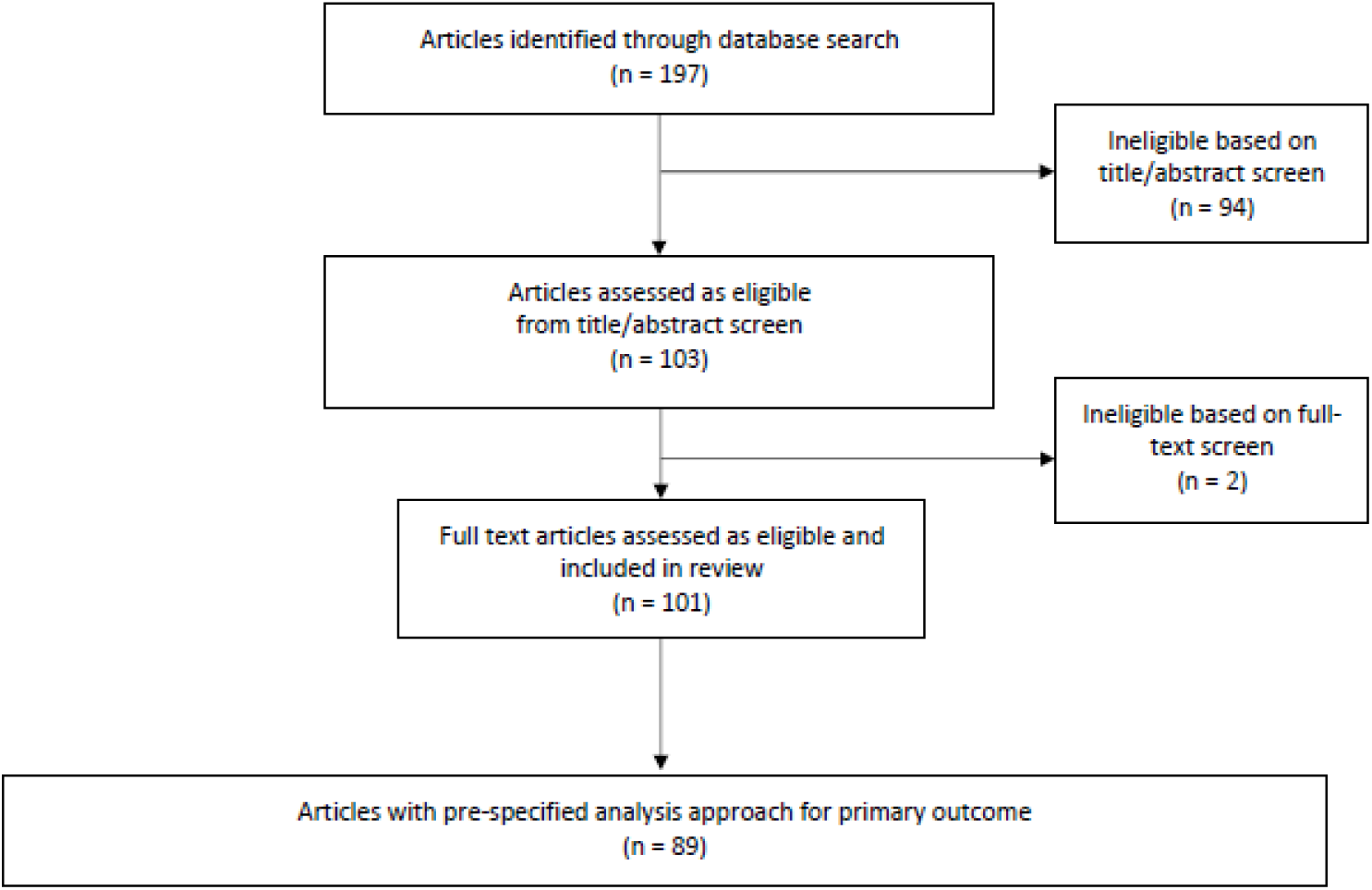
Flow chart of article selection.

Protocols were available for 90 trials (89%) (48 published, 70 as supplementary material with publication, 5 on a website). SAPs were available for 46 trials (46%) (3 published, 43 as supplementary material with publication, 2 on a website). Of 90 trials with an available protocol, the earliest version available was dated before recruitment began for 45 (50%) trials, 19 (21%) were dated during recruitment, 8 (9%) were dated after recruitment ended, and 18 (20%) did not have a date. Of 46 trial with an available SAP, the earliest version of the SAP was dated before recruitment began for 9 (20%) trials, 13 (28%) were dated during recruitment, 13 (28%) were dated after recruitment ended, and 11 (24%) did not have a date.

Overall, only 11 trials (11%) stated in the trial publication, protocol, or SAP that the statistician was blinded until the SAP was signed off and 10 (10%) stated the statistician was blinded until the database was locked.

### Availability of pre-specified analysis approach

Overall, 89 of 101 trials (88%) had a publicly available pre-specified analysis approach for the primary outcome. Eleven trials did not have an available protocol or SAP, and one trial had a protocol with no information on the analysis and no SAP. The document containing the original analysis plan (83 in a protocol, 6 in a SAP) was dated before the start of recruitment for 41 of 89 (46%) trials, during recruitment in 19 (21%) trials (median 19 months post-recruitment beginning, IQR 9 to 46), and after the end of recruitment in 8 (9%) trials (median 7 months post-recruitment completion, IQR 4 to 13). In 21 trials (24%) no date was available.

### Comparison of pre-specified and conducted statistical analysis approach

Of the 89 trials with an available pre-specified analysis approach, only 22 (25%) did not have any unexplained discrepancies (no discrepancies n=5, explained discrepancies only n=17). A further 54 trials (61%) had one or more unexplained discrepancies (see Fig 2). In 13 trials (15%) it was unclear whether an unexplained discrepancy occurred due to poor reporting of statistical methods (unclear whether discrepancy occurred n=11, unclear whether discrepancy explained n=2).

**Fig 2:**
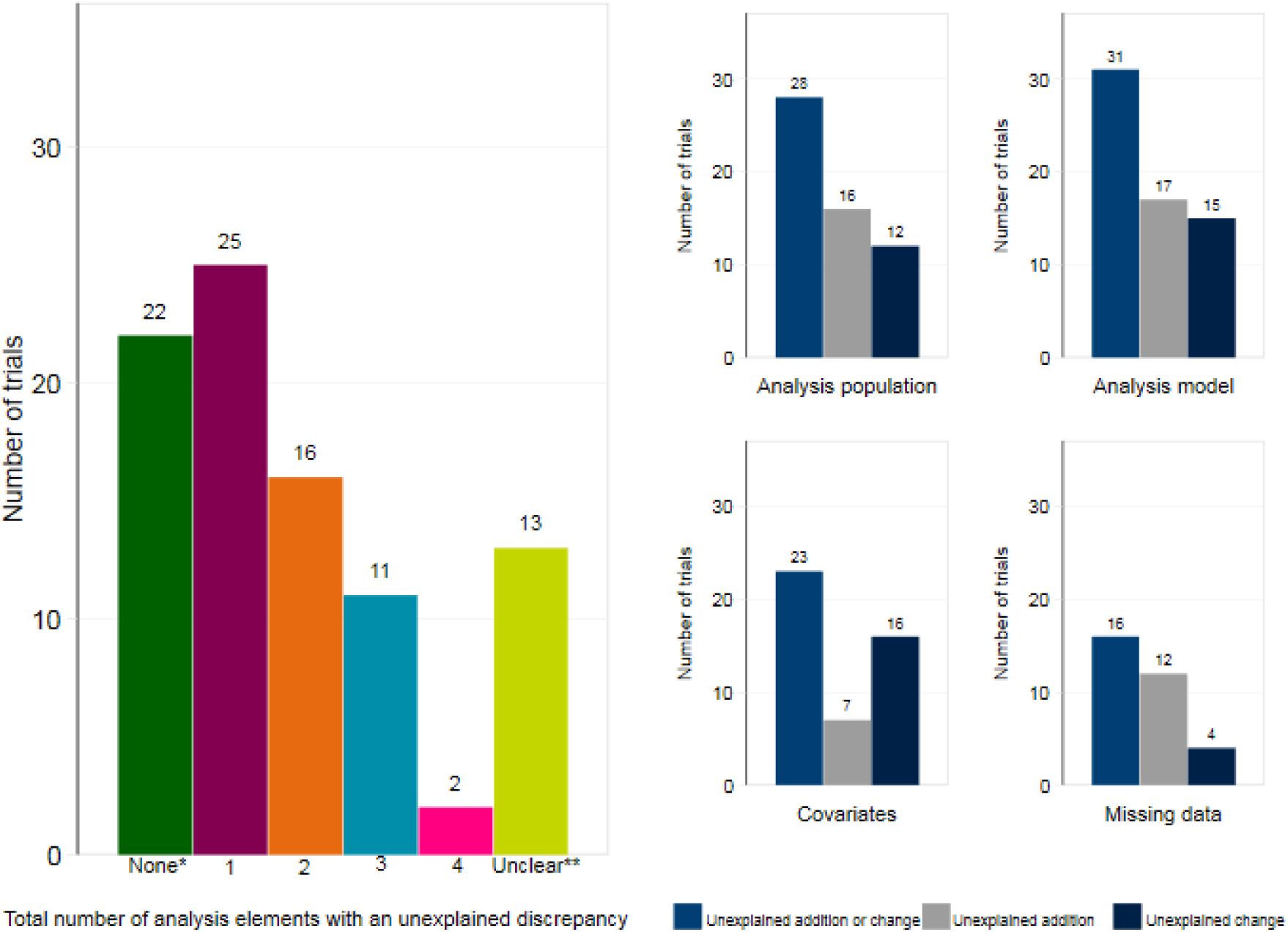
– Number of trials with unexplained discrepancies (Total N=89) ^*^Of the n=22 trials with none; no discrepancies (n=5), explained discrepancies only (n=17). ^**^Unclear if discrepancy occurred (n=11), unclear if discrepancy explained (n=2). One trial had both a change and an addition for the analysis model.

Most trials had one (n=25, 28%) or two (n=16, 18%) unexplained discrepancies. Only 11 (12%) had three and 2 (2%) had four unexplained discrepancies. Unexplained discrepancies were most common for the statistical analysis model (n=31, 35%) and analysis population (n=28, 31%), followed by the use of covariates (n=23, 26%) and handling of missing data (n=16, 18%). Table 2 provides a description of the unexplained discrepancies.

**Table 2.** – Description of unexplained discrepancies (N=89)

| Unexplained changes | N (%) |
| --- | --- |
| Analysis population |  |
| Changed set of patients included by specifying additional exclusions | 12 (13%) |
| Analysis model |  |
| Changed model | 13 (15%) |
| Changed method of selecting analysis model | 2 (2%) |
| Covariates |  |
| Changed from unadjusted to adjusted analysis | 7 (8%) |
| Changed from adjusted to unadjusted analysis | 4 (4%) |
| Changed set of covariates included in analysis | 5 (6%) |
| Missing data |  |
| Changed from complete case to multiple imputation | 1 (1%) |
| Changed imputation strategy | 2 (2%) |
| Unexplained additions |  |
| Analysis population |  |
| Not mentioned in Original Analysis Plan | 8 (9%) |
| Incomplete detail given in Original Analysis Plan | 7 (8%) |
| Allowed analyst to subjectively choose analysis population based on trial dataset | 1 (1%) |
| Analysis model |  |
| Not mentioned in Original Analysis Plan | 5 (6%) |
| Incomplete detail given in Original Analysis Plan | 5 (6%) |
| Allowed analyst to subjectively choose analysis model based on trial dataset | 7 (8%) |
| Covariates |  |
| Not mentioned in Original Analysis Plan | 2 (2%) |
| Incomplete detail given in Original Analysis Plan | 2 (2%) |
| Allowed analyst to subjectively choose covariates based on trial dataset | 3 (3%) |
| Missing data |  |
| Not mentioned in Original Analysis Plan | 9 (10%) |
| Incomplete detail given in Original Analysis Plan | 3 (3%) |
| Allowed analyst to subjectively choose missing data approach based on trial dataset | 1 (1%) |

Overall, 29 trials (33%) had at least one explained discrepancy. Most discrepancies were explained in a later version of the protocol or SAP; only 2 trials explained a discrepancy in the trial publication. Of the 29 trials with an explained discrepancy, only 6 (21%) stated that the statistician was blinded until the SAP was signed off, and 4 (14%) until the database was locked.

#### Subgroup analyses

A total of 43/61 (66%) trials that were not for profit only had at least one unexplained discrepancy, compared to 11/28 (45%) trials that were for profit only. Fewer trials with a SAP available had unexplained discrepancies than trials without an available SAP, though this figure was still high (SAP available 22/46 [48%] with ≥1 unexplained discrepancy vs. no SAP 32/43 [74%]). Trials with a SAP still had a relatively high number of additions to the analysis method, indicating that methods were not being adequately pre-specified within these SAPs (range 7-15% across analysis elements). See Additional File 1, Appendix 3 and 4 for additional results.

## Discussion

In our review of 101 trials published in high impact general medical journals, we found that most had a pre-specified analysis approach for the primary outcome available in either a protocol or SAP. This is essential to allow transparent assessment of whether inappropriate changes were made to the statistical methods. However, most pre-specified statistical analysis approaches were available in a document that was dated after the trial had begun, or had no date available. It is therefore possible that the analysis approach in these documents may have already been changed from the pre-trial version.

Only 25% of trials did not have any unexplained discrepancies between the trial publication and the pre-specified analysis approach, and only 6% had no discrepancies at all. Most trials had at least one unexplained discrepancy (61%), with 32% of trials having two or more. In 15% of trials it was impossible to assess whether there were unexplained discrepancies due to poor reporting of the statistical methods used. Of note, 33% of trials had one or more explained discrepancies; however, less than a quarter of these trials reported that the statistician was blinded to treatment allocation until the analysis plan was finalised or the database was locked. These alterations may therefore have been made based on unblinded trial data, despite being explained. It was also surprising that only two trials explained a discrepancy in the trial publication, despite requirements by the CONSORT (29) statement to do so.

Our results are broadly consistent with previous reviews. Spence *et al (30)* evaluated the availability of protocols and SAPs for trials published in high impact medical journals, and found similar rates of availability. However, the rates of discrepancies we found were generally lower than those previously reported (8, 10, 20, 21). For example, Chan *et al* compared publications to protocols for 70 trials that received ethical approval by the scientific-ethics committees for Copenhagen and Frederiksberg, Denmark in 1994-5 (21). Overall, 44% of trials had unexplained discrepancies in the analysis population, 60% in the analysis model, 82% in the use of covariates, and 80% for handling of missing data. There are several potential explanations for these differences. The introduction of the SPIRIT guidelines in 2013 (25, 26) may have led to better reporting of statistical methods in trial protocols. We also accessed statistical analysis plans in almost half of trials, which increased the number of explained discrepancies. Finally, we evaluated a different population of trials; most of the high impact general medical journals in our review required submission of the trial protocol alongside the article, and may have been less likely to accept trials with extreme discrepancies.

The key issues we identified in this study were: (i) low availability of pre-trial protocols and analysis plans; (ii) poor pre-specification of statistical methods within protocols and analysis plans; (iii) frequent unexplained discrepancies in the final trial publication; (iv) poor reporting of the blinding status of statisticians in relation to modifications of analysis methods or access to trial data; and (v) poor descriptions of the actual analysis methods used in the final publication. Increased adherence to guidelines such as SPIRIT, CONSORT, and the guidelines for Statistical Analysis Plans (6, 26, 27) would help, though alternative approaches to increase transparency around the statistical methods are also required. Two simple proposals that would greatly improve the situation are (a) journals could require authors to submit the first and last version of their protocol and SAP alongside the results article, and publish these as supplementary material; this would allow transparent evaluation of modifications to the analysis approach and be more effective than relying on authors to publish these documents; and (b) journals could require that authors include the statistical code used to perform their analysis alongside the article as supplementary content to allow a complete and transparent comparison of the planned methods vs the final methods (31).

Our study had some limitations. We only included articles from six high impact medical journals; it is likely that trials published in other journals may have lower availability of protocols and SAPs, and higher rates of unexplained discrepancies. Comparisons were based on the first available protocol or SAP, however many were dated after the trial had begun, so there may have been discrepancies before this that we missed.

## Conclusions

In conclusion, unexplained discrepancies in the statistical methods of randomized trials are common. Increased transparency around the statistical methods used in randomized trials is required for proper evaluation of trial results.

## Data Availability

The datasets used and analysed during the current study are available from the corresponding author on reasonable request.

## List of abbreviations

CONSORT: Consolidated Standards of Reporting Trials
GEE: Generalized Estimating Equations
IQR: Interquartile Range
SAP: Statistical Analysis Plan
SPIRIT: Standard Protocol Items: Recommendations for Interventional Trials

## Declarations

### Ethical approval and consent to participate

No ethical approval or consent was required for this review of previously published trials.

### Consent for publication

Not applicable

### Availability of data and materials

The datasets used and/or analysed during the current study are available from the corresponding author on reasonable request.

## Author contributions

The corresponding author affirms that all listed authors meet authorship criteria and that no others meeting the criteria have been omitted. SC and BK had access to all the data in the study and take full responsibility for the work and the conduct of the study and controlled the decision to publish.

Study concept: BK

Study design: BK, SC and GF

Acquisition and interpretation of data: All authors.

Statistical analysis: SC

Drafting of the manuscript: SC and BK

Critical revision of the manuscript for important intellectual content: All authors

## Acknowledgements

Brennan Kahan is grateful for support from the UK Medical Research Council, grant MC_UU_12023/21.

## Additional Material

Additional File 1.doc - Supplementary material – contains additional methods and results. Additional File 2.doc - Protocol and data extraction form – contains protocol and data extraction form used for this study.

## References

1. Prakash A, Risser RC, Mallinckrodt CH. The impact of analytic method on interpretation of outcomes in longitudinal clinical trials. International journal of clinical practice. 2008;62(8):1147–58.

2. Saquib N, Saquib J, Ioannidis JPA. Practices and impact of primary outcome adjustment in randomized controlled trials: meta-epidemiologic study. BMJ : British Medical Journal. 2013;347:f4313.

3. Porta N, Bonet C, Cobo E. Discordance between reported intention-to-treat and per protocol analyses. J Clin Epidemiol. 2007;60(7):663–9.

4. Melander H, Ahlqvist-Rastad J, Meijer G, Beermann B. Evidence b(i)ased medicine--selective reporting from studies sponsored by pharmaceutical industry: review of studies in new drug applications. Bmj. 2003;326(7400):1171–3.

5. Greenberg L, Jairath V, Pearse R, Kahan BC. Pre-specification of statistical analysis approaches in published clinical trial protocols was inadequate. Journal of Clinical Epidemiology. 2018;101:53–60.

6. Gamble C, Krishan A, Stocken D, Lewis S, Juszczak E, Dore C, et al. Guidelines for the Content of Statistical Analysis Plans in Clinical Trials. Jama. 2017;318(23):2337–43.

7. Page MJ, McKenzie JE, Forbes A. Many scenarios exist for selective inclusion and reporting of results in randomized trials and systematic reviews. Journal of Clinical Epidemiology. 2013;66(5):524–37.

8. Li G, Abbade LPF, Nwosu I, Jin Y, Leenus A, Maaz M, et al. A systematic review of comparisons between protocols or registrations and full reports in primary biomedical research. BMC medical research methodology. 2018;18(1):9-.

9. Chan AW, Hrobjartsson A, Haahr MT, Gotzsche PC, Altman DG. Empirical evidence for selective reporting of outcomes in randomized trials: comparison of protocols to published articles. Jama. 2004;291(20):2457–65.

10. Dwan K, Altman DG, Cresswell L, Blundell M, Gamble CL, Williamson PR. Comparison of protocols and registry entries to published reports for randomised controlled trials. Cochrane Database of Systematic Reviews. 2011(1).

11. Chan AW, Altman DG. Identifying outcome reporting bias in randomised trials on PubMed: review of publications and survey of authors. Bmj. 2005;330(7494):753.

12. Dwan K, Altman DG, Arnaiz JA, Bloom J, Chan AW, Cronin E, et al. Systematic review of the empirical evidence of study publication bias and outcome reporting bias. PloS one. 2008;3(8):e3081.

13. Hahn S, Williamson PR, Hutton JL. Investigation of within-study selective reporting in clinical research: follow-up of applications submitted to a local research ethics committee. Journal of evaluation in clinical practice. 2002;8(3):353–9.

14. Ramagopalan S, Skingsley AP, Handunnetthi L, Klingel M, Magnus D, Pakpoor J, et al. Prevalence of primary outcome changes in clinical trials registered on ClinicalTrials.gov: a cross-sectional study. F1000Research. 2014;3:77.

15. Rising K, Bacchetti P, Bero L. Reporting Bias in Drug Trials Submitted to the Food and Drug Administration: Review of Publication and Presentation. PLOS Medicine. 2008;5(11):e217.

16. Vedula SS, Bero L, Scherer RW, Dickersin K. Outcome Reporting in Industry-Sponsored Trials of Gabapentin for Off-Label Use. New England Journal of Medicine. 2009;361(20):1963–71.

17. Williamson PR, Gamble C, Altman DG, Hutton JL. Outcome selection bias in meta-analysis. Statistical methods in medical research. 2005;14(5):515–24.

18. Goldacre B, Drysdale H, Dale A, Milosevic I, Slade E, Hartley P, et al. COMPare: a prospective cohort study correcting and monitoring 58 misreported trials in real time. Trials. 2019;20(1):118.

19. Chen T, Li C, Qin R, Wang Y, Yu D, Dodd J, et al. Comparison of Clinical Trial Changes in Primary Outcome and Reported Intervention Effect Size Between Trial Registration and Publication. JAMA Network Open. 2019;2(7):e197242–e.

20. Dwan K, Altman DG, Clarke M, Gamble C, Higgins JPT, Sterne JAC, et al. Evidence for the Selective Reporting of Analyses and Discrepancies in Clinical Trials: A Systematic Review of Cohort Studies of Clinical Trials. PLOS Medicine. 2014;11(6):e1001666.

21. Chan A-W, Hróbjartsson A, Jørgensen KJ, Gøtzsche PC, Altman DG. Discrepancies in sample size calculations and data analyses reported in randomised trials: comparison of publications with protocols. BMJ. 2008;337:a2299.

22. Editorial Review of Protocols for Clinical Trials. New England Journal of Medicine. 1990;323(19):1355-.

23. Powers JH, Dixon CA, Goldberger MJ. Voriconazole versus Liposomal Amphotericin B in Patients with Neutropenia and Persistent Fever. New England Journal of Medicine. 2002;346(4):289–90.

24. International Council for Harmonisation of Technical Requirements for Pharmaceuticals for Human Use. ICH Harmonised Tripartite Guideline: Statistical Principles for Clinical Trials E9.: London, England: European Medicines Agency; 1998.

25. Chan AW, Tetzlaff JM, Altman DG, Laupacis A, Gotzsche PC, Krleza-Jeric K, et al. SPIRIT 2013 statement: defining standard protocol items for clinical trials. Ann Intern Med. 2013;158(3):200–7.

26. Chan A-W, Tetzlaff JM, Gøtzsche PC, Altman DG, Mann H, Berlin JA, et al. SPIRIT 2013 explanation and elaboration: guidance for protocols of clinical trials. BMJ : British Medical Journal. 2013;346:e7586.

27. Moher D, Hopewell S, Schulz KF, Montori V, Gøtzsche PC, Devereaux PJ, et al. CONSORT 2010 Explanation and Elaboration: updated guidelines for reporting parallel group randomised trials. BMJ. 2010;340:c869.

28. StataCorp. Stata Statistical Software: Release 15. College Station, TX: Stata Corp LLC. 2017.

29. Altman DG, Schulz KF, Moher D, Egger M, Davidoff F, Elbourne D, et al. The revised CONSORT statement for reporting randomized trials: explanation and elaboration. Ann Intern Med. 2001;134(8):663–94.

30. Spence OM, Hong K, Onwuchekwa Uba R, Doshi P. Availability of study protocols for randomized trials published in high-impact medical journals: A cross-sectional analysis. Clinical Trials. 0(0):1740774519868310.

31. Goldacre B, Morton CE, DeVito NJ. Why researchers should share their analytic code. BMJ. 2019;367:6365.

